# Socio-economic and Demographic Factors Influencing Interpersonal Communication Between Patients and Family Physicians: A Protocol for a Systematic Review

**DOI:** 10.1101/2023.09.18.23295671

**Authors:** Olha Svyntozelska, Nataly R. Espinoza Suarez, Juliette Demers, Michèle Dugas, Annie LeBlanc

## Abstract

**Background:** Interpersonal communication is an essential aspect of patients’ relationships with their family physicians. It impacts patients’ experiences and the quality of care. Nevertheless, interpersonal communication can be affected by conscious and unconscious biases based on patients’ socio-economic and demographic characteristics. Evidence synthesis of this aspect of interpersonal communication in primary health care is limited. This systematic review will assess socio-economic and demographic factors influencing interpersonal communication between family physicians and patients living with one or more chronic conditions during clinical encounters.

**Methods:** We will perform a systematic review following the Joanna Briggs Institute Manual for Evidence Synthesis. The population of interest is adults living with at least one chronic condition. We will collect socioeconomic and demographic factors such as gender, sex, race or ethnicity, levels of literacy and/or health knowledge, level of education, and poverty or socioeconomic status, including employment or income level. Any published empirical study reporting aspects of interpersonal communication between patients and their family physicians will be considered. Three databases (Embase, MEDLINE, and Cochrane) will be assessed for eligible studies. Pairs of independent reviewers will select studies, extract data, and conduct bias assessment using MMAT-2018. We anticipate conducting descriptive and content analyses with narrative synthesis.

**Discussion:** Findings from this review may guide better communication between primary care physicians and their patients and increase awareness of potential health inequalities pathways in clinical practice.

**Registration number:** CRD42023411895 (PROSPERO platform).

## INTRODUCTION

Primary healthcare is a multilevel structure at the heart of health systems and is responsible for continuously supporting patients throughout their life journey (1). Patient-centered care is replacing the disease-centered care model across primary health care, as it takes into consideration patients’ needs and desires while coordinating the integration of services and care provided by other specialties (2, 3).

Interpersonal communication is both an important dimension of primary healthcare and an essential component of clinician-patient interactions (4-6). Interpersonal communication is defined as the clinician’s ability to elicit and understand patients’ concerns, explain healthcare options, and engage patients in shared decision-making (4, 5). Its most important aspects are verbal communication (e.g., use of open-ended questions, level of language) and nonverbal communication (e.g., expression of empathy, type of eye contact) (4, 5). Together, they contribute to relationship building, information gathering and sharing, decision-making, expressing empathy, and encouraging disease self-management (7).

However, interpersonal communication can be completely or partially influenced by clinicians’ personal biases, whether conscious or not (8, 9). These biases may disadvantage groups of people who are vulnerable or socially disadvantaged due to their socioeconomic status, including employment and/or income level, ethnic or socio-cultural background, gender or sex, education level, and literacy and/or health literacy (10, 11).

These biases influencing interpersonal communication have been documented to impact different aspects of care. Patients from lower socioeconomic statuses perceived clinicians spent less time with them during consultations and were less attentive and less empathetic, which negatively affected their perception of the information-sharing and shared decision-making processes (12). Patients’ ethnic backgrounds have been shown to impact the quality of interpersonal communication as well as treatment strategies (9, 13). Impacts on interpersonal communication have equally negative consequences for patients’ disease management (5, 14). The importance of interpersonal communication improvement has been shown for outcomes such as hospitalizations, symptom prevalence, and adherence to the therapy of choice (15).

Almost half of Canadians aged 20 years and older live with one or more chronic conditions, and its prevalence is known to increase, especially among disadvantaged population groups (16, 17). Thus, it is important to acknowledge disadvantaged population groups in the primary care of adults living with chronic conditions, as their mere presence in interpersonal communication with their physician can have an immeasurable societal and personal impact (18).

The most recent literature review examining socio-economic differences in patient-clinician communication was conducted in 2012 and was limited to the concepts of education level, occupation, and income (8). To improve the understanding of this phenomenon after ten years of ongoing research, updating existing knowledge with the enlargement of review to other socioeconomic and demographic factors seems indispensable. Thus, we aim to assess socio-economic and demographic factors influencing interpersonal communication between family physicians and patients living with one or more chronic conditions during clinical encounters.

## METHODOLOGY

We will conduct a systematic review in accordance with the Joanna Briggs Institute Manual for Evidence Synthesis and will follow the Preferred Reporting Items for Systematic Reviews and Meta-Analyses guidelines (PRISMA) for its reporting (19, 20). This review is registered on the PROSPERO platform (CRD42023411895).

### Eligibility Criteria

We followed the framework Population, Intervention/Exposure, Comparison, and Outcomes (PICO) in establishing our eligibility criteria (21).

#### Population

We will include studies of adults (18 years and older) living with one or more physical chronic conditions (e.g., diabetes, high blood pressure, chronic bronchitis/chronic obstructive pulmonary disease), regardless of gender, ethnicity, religion, or geographic area. We will also include studies of individuals living with both mental health disorders and chronic conditions. Studies reporting findings for adults with only mental health issues will be excluded.

#### Exposure

We will include studies focusing on socio-economic and demographic factors. More specifically, we will include studies of individuals in disadvantaged socio-economic or demographic groups as per previously published definitions: (i) socially disadvantaged due to poverty or lower socio-economic status including employment or income level; (ii) socially disadvantaged due to their ethnic or socio-cultural background; (iii) gender or sex; (iv) lower level of education (no university degree); (v) low levels of literacy and/or health knowledge (10, 11).

#### Outcomes of interest

The outcome of interest is interpersonal communication, reported across the following components: verbal dominance of the conversation by the physician, attentive behaviour (physician actively listens), use of plain language by the physician, maintenance of eye contact by the physician, expression of empathy, tone of voice, body posture, encouragement of shared decision-making (7, 22).

#### Study setting

We will include studies of encounters in primary care clinical settings. We will exclude studies of visits to other health services that do not include family physicians (e.g., visits to pharmacists, social workers, and nurses as well as specialized care units).

#### Type of design and materials

We will include mixed, qualitative, and quantitative observational (e.g., cohort, cross-sectional, case-control) or experimental peer-reviewed studies. Systematic reviews will be excluded, but we will consider them as sources of data for additional searches of relevant primary studies. Case series and case reports, as well as commentaries, letters to the editor and others, theses, conference abstracts, and annual reports or research reports will be excluded.

We will not apply any restrictions on the language or year of publication.

### Information Sources and Search Strategies

The search strategy will be developed under the leadership of an information specialist. The search will be conducted in Embase, MEDLINE (via PubMed), and Cochrane. An initial search will be developed for MEDLINE, with a later adaptation of syntaxis to match other information sources. A mix of controlled (e.g., “Socioeconomic Factors”[Mesh], “Healthcare Disparities”[Mesh]) and free vocabulary (e.g., Socio-economic Factors [TIAB], Healthcare Disparities [TIAB]) was used to search for the main concepts. No review of grey literature is planned.

### Selection, Extraction and Management of Data

#### Selection process

Pairs of reviewers will conduct the pilot screening with the use of distributed criteria for article selection. After, they will continue independently onto the screening by titles and abstracts, followed by full-text screening and data extraction. Disagreements will be resolved by discussion and involvement of a senior reviewer if needed.

#### Data collection process

Extracted information will include: (1) study identification information (e.g., title, first author, year of publication); (2) study characteristics (e.g., country, study design, sample size); (3) population of the study information (e.g., age, sex and gender, ethnic background); (4) socio-economic and demographic factors of interest in the study, their definitions, and tools used to measure them; (5) reported outcomes (e.g., empathy, tone of voice, body posture of a physician), their definitions, and tools used for the collection; (6) effect measures when available.

#### Quality assessment

Risk of bias and quality assessment will be conducted by pairs of reviewers independently with the use of MMAT, a tool well adapted for different study designs (23). This will give us uniformity in the assessment of both qualitative, quantitative, and mixed studies. Any arising discrepancy will be discussed by the reviewers until a consensus is reached.

#### Data management

EndNote 20 (Clarivate) will be used to store references, while DistillerSR (DistillerSR. V.2.35. Evidence Partners; 2023. Accessed January 2023–February 2023. https://www.distillersr.com) will be the main tool in the article selection and extraction process as well as in the calculation of the discordance statistics.

### Data Synthesis and Analysis

We will synthesize data on the risk factors influencing different communication behaviours in the clinical encounter as well as their potential effects. Particular attention will be paid to the differences in the tools used to measure communication behaviours and risk factors. We will also explore different definitions used in studies to both define communication behaviours and their risk factors. Due to the anticipated heterogeneity of studies, no meta-analysis is planned.

### Patient and Public Involvement

Preliminary results will be discussed with patients to improve interpretations and understanding of possible implications. Results will be distributed amongst direct and indirect stakeholders as well as disseminated via scientific conferences, research webinars, and publications in peer-reviewed journals.

### Ethics and Dissemination

Due to the absence of human or animal subjects’ involvement, no ethical approval is necessary for the systematic review.

## DISCUSSION

Although interpersonal communication is a key component of clinical encounters, existing systematic reviews need to be broadened in terms of socio-economic and demographic factors that might impact patient-physician communication. With the possible surge of research focusing on socio-economic and demographic determinants of health, one of the core dimensions of primary healthcare, patient-physician communication needs to be re-evaluated through the prism of socio-economic and demographic influences. Thus, this systematic review has the potential to address and further our understanding of what factors might play a role in interpersonal communication during clinical encounters as well as how these factors influence interpersonal communication as per current knowledge.

## Data Availability

All data utilized in the present study are available upon reasonable request to the authors.

